# YouTube as an information source during the Coronavirus disease (COVID-19) pandemic: Evaluation of the Turkish and English content

**DOI:** 10.1101/2020.05.06.20093468

**Authors:** Ömer Ataç, Yunus Can Özalp, Rifat Kurnaz, Osman Murat Güler, Melikşah İnamlık, Osman Hayran

**Affiliations:** Department of Public Health, School of Medicine, Istanbul Medipol University; School of Medicine, Istanbul Medipol University

**Keywords:** SARS-CoV-2, DISCERN, MICI, Video Power Index

## Abstract

**Bakcground:** YouTube is an important online source of information. And its viewing numbers tend to increase exponentially in extraordinary situations. Our aim in this study was to evaluate the contents of the most frequently viewed YouTube videos during the COVID-19 pandemic.

**Methods:** In this study, contents of the most frequently viewed Turkish and English videos regarding COVID-19 pandemics are examined and scored with modified DISCERN, MICI and VPI.

**Results:** The mean DISCERN score of Turkish videos is similar to English videos (2.55±1.40 and 2.43±1.25 respectively). Total MICI score tends to be higher in Turkish videos. 86.9% of all 168 videos and 65.2% of all 23 misleading videos were released by news channels. Average view counts, view ratios, and VPIs of misleading videos are higher than the useful videos.

**Discussion:** Since there is not a peer-review system on YouTube, it is very important for the content of videos that are released through news channels to be accurate because the important messages can be spread among people in society through them. Especially some Turkish videos included many different rumors and faulty statements. During the extraordinary situations such as the pandemics, the videos of official health authorities and international institutions should be more visible in YouTube.

## 1. Introduction

COVID-19 was primarily announced to the world with unknown etiology pneumonia cases from the city of Wuhan where is located at the state of Hubei at China [1]. The situation report of the World Health Organization (WHO), which was published on 30 January 2020 described it as interim name Novel Coronavirus (2019-nCov) [2]. On 5 February 2020, the name of the virus was announced as SARS-CoV-2 by The International Committee on Taxonomy of Viruses (ICTV) and the disease that the virus leads to was called COVID-19 in a press conference of the WHO Director General [3-5]. While the outbreak of the disease was initially identified as Public Health Emergency of International Concern on January 30, it was later announced as pandemic on March 11 [6,7]. By the first week of April, the confirmed cases exceeded 1 million and reached to 212 different territories [8]. As of April 18, the number of confirmed cases and death were 2.160.207 and 146.088, respectively [9]. The first confirmed case at Turkey was first seen on March 10 and the first death happened on March 15. As of April 18, the number of confirmed cases and death are 82.329 and 1890, respectively [10].

The virus spread among humans at close distance through droplet. The other common type of contagion is the spread through mucosa with contaminated hands [11]. Transmission ability of asymptomatic cases is one of the difficulties in pandemic. Since there is no vaccine yet, personal protective measures such as hand-washing, physical and social distancing, and isolation of the infected individuals are the most effective ways to struggle against pandemic [11]. People need to access correct information and then apply them in their lives to achieve this. Study results indicate that social media and online platforms in the internet are the major sources of medical information for many people [12,13]. It is important to access correct and reliable through these channels. However, almost none of these platforms and social media are peer-rewieved and they may include many false or misleading information.

YouTube is an important online source of information with its 2 billion users globally. Its viewing numbers tend to increase exponentially in extraordinary global situations [14]. Not only ordinary people or patients, but healthcare institutions and professionals also use and share information via YouTube [15]. Because of the open access and lack of peer review, there are some concerns related with reliability, confidentiality and privacy of contents [16,17]. It is suitable for misinformation, disinformation and anecdotes which are not based on any evidence [13,18]. In the literature regarding the past pandemics, it was demonstrated that the percentage of videos which contain false information is between 8.0% and 23.8% on YouTube [19–21].

The content analysis regarding the social media and online platforms is a significant research issue in recent years. The nature of the health information spread through internet is crucial especially during extraordinary times such as disease pandemics and increasingly investigated [20,22,23]. Our aim in this study was to review and evaluate the contents of the most frequently viewed YouTube videos during the COVID-19 pandemic.

## 2. Materials and methods

This study is conducted as a qualitative study. Contents of the most frequently viewed Turkish and English YouTube videos regarding COVID-19 pandemics are examined during April 2020.

### 2.1. Selection of the study material

On April 9 2020, the search process was conducted on YouTube by using both Turkish and English keywords such as ‘’Corona virüsü’’, ‘’Koronavirüs’’, and ‘’Koronavirüs Hastalığı’’; ‘’COVID-19’’ and ‘’Corona virus’’. In order to prevent the influence of cache, cookies, and watch history on the search process, a new YouTube account was created for this study. In all searches, the relevancy level in filter was selected as default on YouTube. The first 50 results were recorded in separate list based on each keyword. The reason why the first 50 results were selected is that some studies show that YouTube users do not tend to watch videos after a couple of pages [24]. These videos are reviewed and examined in accordance with the stages in Figure 1. Since the literature demonstrates that the optimal length for a YouTube video is between 10 and 16 minutes, those who exceeded the 15 minute-threshold were eliminated during the study [25]. Ultimately, 101 Turkish and 67 English videos, which met these criteria, were included to the study. Since this study is conducted through open data that are accessible to all people, any ethics committee approval was not taken.

**Figure 1:**
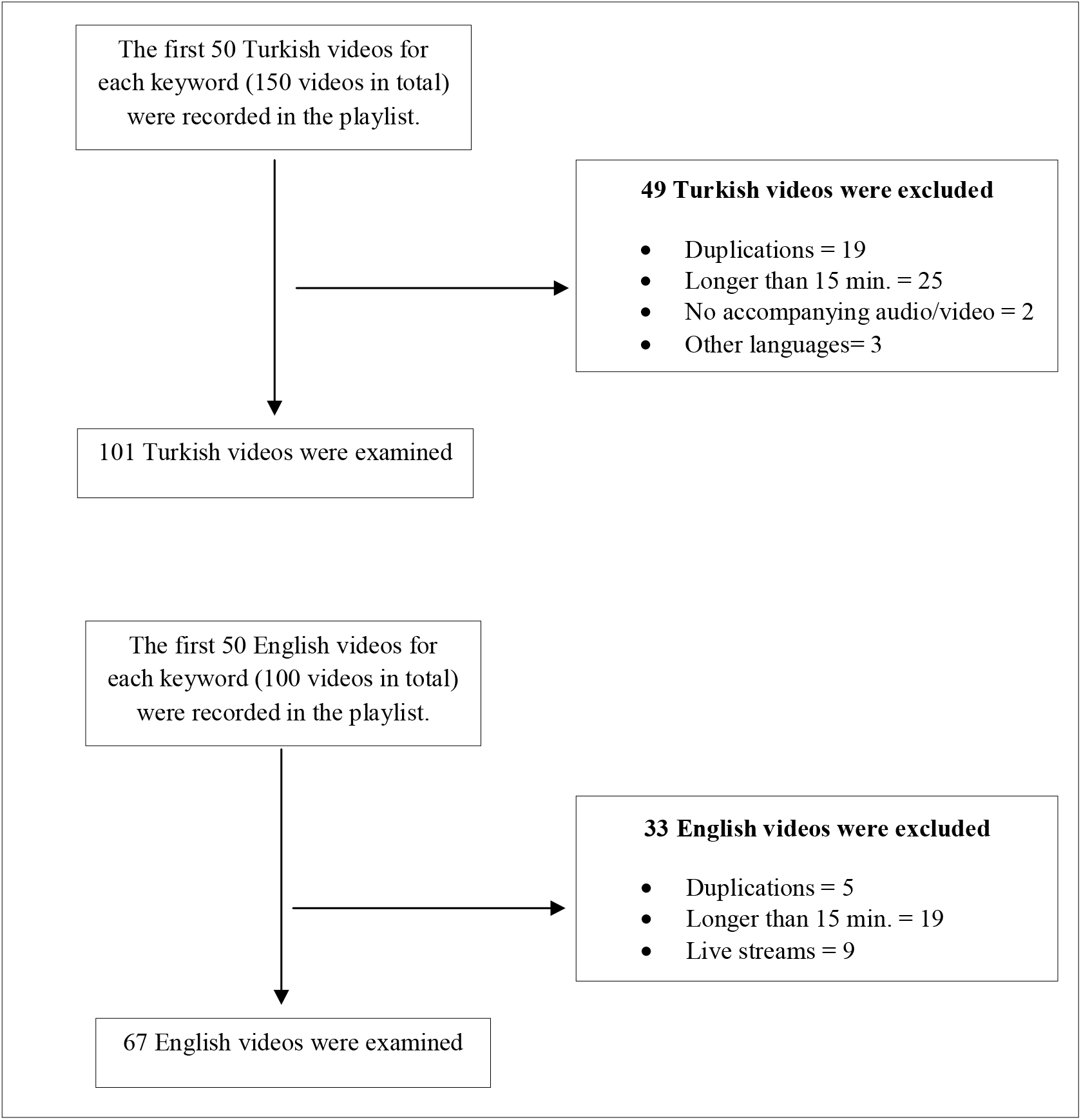
PRISMA Flow Diagram for selection process

### 2.2. Evaluation of the contents

The descriptive characteristics such as the name of videos, their upload dates, view counts, likes, sources and content were recorded on April 10 2020. Every video was evaluated through the principles of DISCERN and medical information and content index (MICI). Moreover, a Video Power Index (VPI) was calculated for each video and the evaluations of modified DISCERN and MICI were conducted by researchers [20,22,26,27].

**Table 1:** Modified DISCERN

|  |
| --- |
| 1. Are the aims clear and achieved? |
| 2. Are reliable sources of information used? (i.e., publication cited, speaker is a certified physician) |
| 3. Is the information presented balanced and unbiased? |
| 4. Are additional sources of information listed for patient reference? |
| 5. Are areas of uncertainty mentioned? |

Modified DISCERN is a five-question scale that was adapted by Singh from a sixteen-question DISCERN tool which was developed by Charnock et al [27,28]. Each criterion is ranked as 1-0 (yes/no) and scored between 0 and 5.

MICI was developed by Nagpal et al. during the period of Ebola epidemic in order to evaluate the content quality of videos that contain medical information and has been used by studies about the COVID-19 [20,22]. It examines every video under these main categories: prevalence, transmission, signs and symptoms, screening/testing, and treatment/outcome. Each main category includes 5 different criteria which means that there are 25 different criteria in MICI. Every criterion is ranked as 1-0 and scored between 0 and 25 (Supplement-1).

VPI ([(view ratio x like ratio/100]), where view ratio=views/day and like ratio=[(likes x 100)/ (likes + dislikes)] was developed as an index by Erdem in order to measure the power of social media based on the descriptive features of videos and has been used by different studies [12,26].

While two researchers (Mİ and YCÖ) evaluated the Turkish videos, other two researchers (RK and OMG) examined the English videos separately for eligibility. A third researcher (ÖA) consulted during the evaluation process when there was a conflicting issue to finalize the decision. Level of agreement between researchers was significantly high for both languages (Cohen’s kappa: 0.81 for Turkish, and 0.85 for English).

The content evaluation was conducted under three categories: useful, misleading, and news update [20,21,29]. Those videos which contain scientific and reliable information are coded as useful while the ones which include false information, conspiracy theory or manipulation are coded as misleading and those which share information through new channels are coded as news update.

### 2.3. Statistical Analysis

After the data was coded in Microsoft Office 365 Excel, it was transferred to SPSS 24.0 for analyses. Mean, standard deviation, frequencies and percentages are calculated for descriptive statistics. Unpaired T-test was used to analyze the differences and the statistically significant level was accepted as p < 0.05.

## 3. Results

The total view count of all 168 videos is 67.222.756. As it is shown in Table 2, the time interval between the release date of English videos and the date of this research is shorter than the case of Turkish videos (p < 0.001). The views/day ratio for English videos is significantly higher than Turkish ones (p < 0.001). Number of comments, length of duration and VPI scores of the English videos are significantly higher than the Turkish videos while the mean DISCERN score of Turkish videos is similar to English videos (2.55±1.40 and 2.43±1.25 respectively).

**Table 2:** Descriptive characteristics of Turkish and English videos

| **Descriptive characteristics** | **Turkish (n=101)**<br>**mean±SD** | **English (n=67)**<br>**mean±SD** | **p** |
| --- | --- | --- | --- |
| **Days since release date** | 28.38±22.78 | 4.22±7.66 | **<0.001** |
| **Number of views** | 373,496.83±673,947.92 | 511,508.78±635,301.38 | 0.079 |
| **Views/day** | 15,606.50±25,034.16 | 194,348.33±307,960.44 | **<0.001** |
| **Number of likes** | 12,764.68±33,054.33 | 6923,89±21.825,83 | 0.183 |
| **Number of dislikes** | 463.42±2061.24 | 816.89±1235.28 | 0.214 |
| **Number of comments** | 689.38±2284.22 | 2641.29±3200.62 | **<0.001** |
| **Length of duration (sec)** | 227.87±180.94 | 359.94±212.35 | **<0.001** |
| **VPI score** | 93.24±198.09 | 1603.19±2304.32 | **<0.001** |
| **DISCERN score** | 2.55±1.40 | 2.43±1.25 | 0.556 |

Mean MICI scores for Turkish and English videos are presented in Table 3. Total MICI score tends to be higher in Turkish videos however there is no significant difference between two languages (p=0.212). Mean scores for Transmission and Screening/Testing of Turkish videos are significantly higher than the English videos (p=0.046 and p < 0.001 respectively).

**Table 3:** MICI scores by language

| | **Turkish**<br>mean±SD | **English**<br>mean±SD | p |
| --- | --- | --- | --- |
| **Prevalence** | 0.66±1.44 | 1.07±1.31 | 0.062 |
| **Transmission** | 1.03±1.40 | 0.61±1.18 | **0.046** |
| **Signs-Symptoms** | 1.11±1.64 | 0.24±0.78 | **<0.001** |
| **Screening/Testing** | 0.11± 0.31 | 0.16±0.73 | 0.501 |
| **Treatment/Outcome** | 0.42±0.87 | 0.67±1.11 | 0.114 |
| **Total MICI Score** | 3.33±3.09 | 2.76±2.49 | 0.212 |

86.9% of all 168 videos were released in news channels (Table 4). 33.6% of them are categorized as useful. 15.8% of Turkish videos and 10.4% of English videos have a misleading content. 65.2% of all 23 misleading videos were released by news channels.

**Table 4:** Distribution of videos by language, source of release and content

| **Source of release** | **Turkish (n=101)** |  |  | **English (n=67)** |  |  |
| --- | --- | --- | --- | --- | --- | --- |
|  | Useful (%) | Misleading (%) | News Update (%) | Useful (%) | Misleading (%) | News Update (%) |
| **Ministry/academic/hospital (n=4)** | 4 (8.7) | 0 | 0 | 0 | 0 | 0 |
| **News channels (n=146)** | 35 (76.1) | 9 (56.3) | 39 (100.0) | 14 (82.4) | 6 (85.7) | 43 (100.0) |
| **Other** (n=18) | 7 (15.2) | 7 (43.7) | 0 | 3 (17.6) | 1 (14.3) | 0 |
| **Total** | 46 (100.0) | 16 (100.0) | 39 (100.0) | 17 (100.0) | 7 (100.0) | 43 (100.0) |

When the descriptive characteristics of videos are compared in terms of their content category, it was found out that the average view counts, view ratios, and VPIs of misleading videos are higher than the useful videos, but the difference between these groups is not statistically significant (Table 5). Only likes ratio of useful videos are higher than the misleading videos.

**Table 5:**
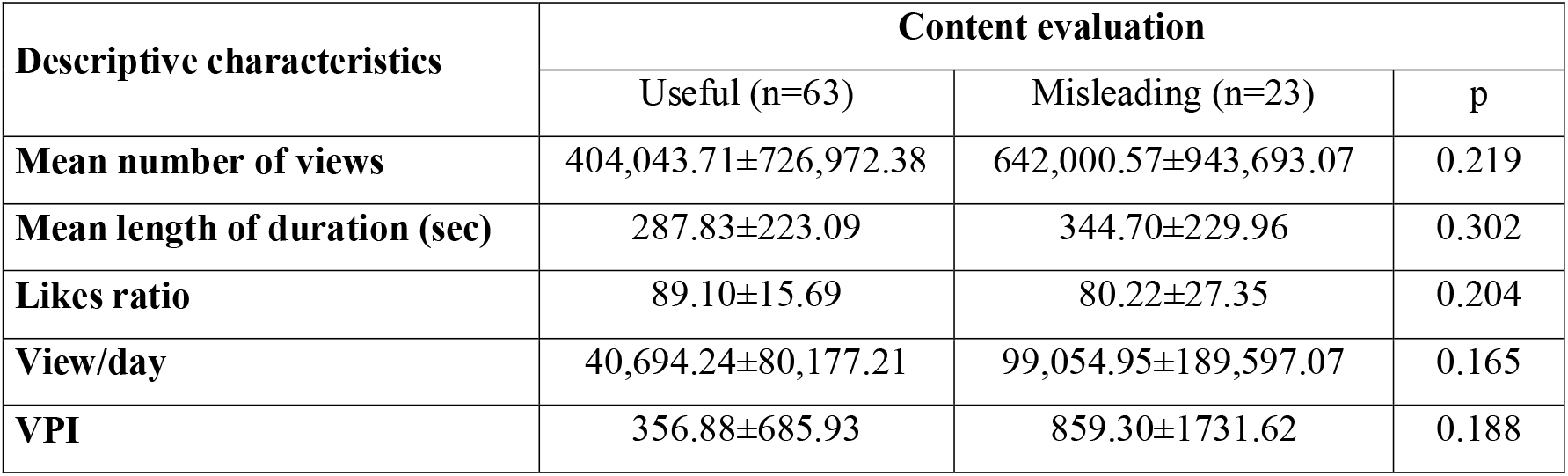
Content evaluation of videos by descriptive characteristics

## 4. Discussion

Our study findings indicate that only 37.5% of the reviewed videos have useful content. A recent study which examined videos in English and Chinese regarding COVID-19 pandemic found the proportion of useful contents as 58.8% [20]. In another study about H1N1 influenza, the proportion of useful video contents were found to be 61.3% [21]. Both studies have found higher proportions of useful contents than our study. The reason for low proportion of useful contents could be the high proportion of misleading content in Turkish videos than English videos (15.8% and 10.4% respectively) in our study group. On the otherhand 65.2% of misleading videos were released by news channels.

In both languages, majority of the videos (86.3%) were released by the news agencies. There was no video from institutions such as WHO, Centers for Disease Control and Prevention (CDC) and European Centre for Disease Control and Prevention (ECDC) in our study. While Khatri et al.’s study included only 6 (5.3%) videos from the WHO, other study conducted by Pathak et al. did not contain any video from the WHO and CDC [20,29]. Four Turkish videos, one by Turkish Ministry of Health, one by an academic institution and two by hospitals, were released by professional health institutions. This finding can be concluded as a lack of interest of the professional health institutions to electronic media.

The difference between the mean DISCERN scores of Turkish and English videos are not statistically significant. The mean scores found by Khatri and his colleagues are higher than our findings [20]. In the results of MICI, the average score of Turkish videos are higher than English videos. In Khatri and his colleagues’ study, both their average score of English and Chinese videos are higher compared to our study results [20]. The reason for the differences in the scores of DISCERN and MICI between our study and Khatri et al.’s study can be that there might be a change in the ranks of English videos from their study time to ours. On the other hand, there might be a bias of researchers in terms of their evaluations and rankings. Nevertheless, it is still significant to see that only six videos had a score of 10 or above in a ranking system that examines the information quality of contents in which the maximum score is 25. Furthermore, four of those videos were released through news channels and none of them was produced by an academic institution or the Ministry of Health. During the COVID-19 period, one research studied the most popular 100 videos in English to examine the information quality of preventive behaviors and indicated only one-third of those videos included one of the seven different preventive behaviors. Moreover, it found out that 79.0% of all videos had a content that can trigger fear and anxiety in the society [23]. In our study, especially some Turkish videos included many different rumors and faulty statements such as; ‘’the virus is revealed in a laboratory environment, its treatment is certain, but they’re waiting for right time to announce it’’. We noticed that even some medical doctors expressed misleading or faulty comments such as ‘’number of cases will not increase”, “it was a virus that should not be feared”, “saltwater mouthwash, vinegar water, or kelle paca soup (a traditional dish consists of a sheep’s head and trotters) prevents this disease”, or even “the virus did not exist at all’’.

There are some limitations in our study. Since this study was conducted in a specific time period, it can lead to different results if it is conducted in different periods because the content and definitive features of YouTube are subject to change instantly. However, since this study was conducted more than three months after the announcement of the first confirmed case, the videos which were analyzed might relatively have a standard ranking. Although kappa coefficient was used for the measurement of DISCERN and MICI scores, there might be some issues regarding both intra- and inter-observer bias in our study. Lastly, it might be also a limitation that we only included videos in two different languages with five different keywords which led us to evaluate 50 videos for each keyword.

YouTube is one of the most common news and information source in today’s world because of its simple access and provision of various contents. However, since there is not a peer-review system on YouTube, except copyright and common complaint issues, people can almost release every type of videos. Consequently, it also becomes a suitable platform for the spread of misinformation and disinformation. News channels are the most-used sources of videos for users. It is very important for the content of videos that are released through these news channels to be accurate because the important messages can be spread among people in society through them. However, the fact that the videos created by international institutions, academic and ministry accounts tend to be watched less than news channels shows that these institutions are not successful in using such platforms. Different solutions should be developed in order to increase the view counts of these institutions. During the extraordinary situations such as the pandemics, the videos of official health authorities and international institutions should be more visible in YouTube. In Turkey, the YouTube videos regarding the COVID-19 are provided through COVID-19 health portals and these portals are linked to the information address of the Ministry of Health (covid19.saglik.gov.tr) [30].

## Data Availability

We declared that all data referred to in the manuscript obtained from YouTube

